# Number of previous caesarean deliveries, history of vaginal birth and uterine rupture during a trial of labour : Protocol for a retrospective population-based cohort study

**DOI:** 10.64898/2026.07.19.26358407

**Authors:** R Thompson, P Adily, M Lauer, R Narayan, A Mackie, H Phipps, V Berghella, B de Vries

## Abstract

There remains considerable uncertainty around the safety of a trial of labour after more than one caesarean delivery. This large retrospective cohort study will investigate the safety of a trial of labour using a large United States dataset of all registered births from 2011 to the most recent year with available data. A multivariable fitted model will be used to predict the probability of uterine rupture in women with two or more previous caesarean deliveries, with a minimum 21-month interpregnancy interval. This will provide important information to clinicians in counselling women wanting to attempt a vaginal birth after more than one caesarean delivery.

## Introduction

The rising rate of caesarean delivery continues to be a public health concern, both in Australia and internationally. Between 2011 and 2023, the rate of caesarean delivery in Australia rose from 32% to 41%^1^. Caesarean deliveries carry significant short- and long-term risks to maternal health. In particular, the rising risk of future pregnancy complications such as placenta accreta spectrum disorder, caesarean scar niches and caesarean scar ectopic pregnancies, present significant challenges in maternal health care^2^.

A number of maternal demographic and clinical factors have been shown to contribute to the rise in rates of caesarean delivery over the last few decades,^3^ however history of previous caesarean delivery continues to be the most common indication for a caesarean birth in the current pregnancy. Most women who have a trial of labour after caesarean (TOLAC) will achieve a vaginal birth^4^. However, a TOLAC is often not attempted, and only 11% of women in Australia with one previous caesarean delivery have a vaginal birth in a subsequent pregnancy^5^. Increasing the rate of VBAC is one potential way to address the rising rates of caesarean deliveries.

One of the most important considerations in the decision to have a TOLAC is the risk of uterine rupture. In many parts of the world including Australia, clinicians will often advise against TOLAC for women with a history of more than one caesarean delivery, as the risk of uterine rupture is considered to be too high^6^. In a re-analysis of the landmark prospective study by Landon et al, no difference was observed in the risk of uterine rupture with multiple compared with one previous caesarean delivery^7^. However, even in this large study, there were only nine uterine ruptures in the group with multiple previous caesarean births. In a subsequent meta-analysis, the combined rate of rupture amongst women having a TOLAC after two caesarean deliveries was 1.4%^8^.

This study will aim to evaluate the safety of TOLAC after two or more caesarean deliveries using a large US population-based data set, by assessing the relationship between number of previous caesareans and the presence of a previous vaginal birth, with the rate of uterine rupture in labour.

### Study design

A retrospective cohort study assessing the relationship between the number of previous caesarean deliveries, previous vaginal deliveries, and the risk of uterine rupture.

### Study setting

The study will include all eligible births in the United States using the 2003 Revision of the US Certificate of Live Birth, from the year 2011 to the most recent year for which data are available (currently 2024).

### Participants

#### Eligibility criteria

- Had a trial of labour
- 17-43 weeks gestational age^
- One to three previous caesarean deliveries^
- Minimum 21-month interpregnancy interval*

^From 17 weeks gestational age because that is the available data and >43 weeks gestational age is excluded due to concerns about misclassification. More than three previous caesareans is also excluded due to concerns about misclassification.

*We previously observed no association between interpregnancy interval and uterine rupture among women with one previous caesarean and no previous vaginal births for intervals longer than 21 months^ix^. When there is a history of both caesarean and vaginal births, the interval from the most recent birth is reported and whether it was a caesarean or vaginal birth is unknown. This eligibility criterion ensures all intervals from the last caesarean to the onset of the current pregnancy are >21 months, and hence the estimates of risk are not influenced by the known association between interpregnancy intervals <21 months and uterine rupture.

#### Exclusion criteria

- Caesarean delivery without a trial of labour
- Fetal congenital anomaly where reported (anencephaly, meningomyelocele/spina bifida, cyanotic congenital heart disease, congenital diaphragmatic hernia, omphalocele, gastroschisis, limb reduction defect, cleft lip/palate, Down Syndrome, suspected chromosomal disorder, hypospadias)
- Multiple pregnancy (excluded because in our previous study, the rate of uterine rupture was very low in twin pregnancies)^9^.
- Non-cephalic presentation
- More than 3 previous caesarean deliveries
- 2003 Revision of the US Certificate of Live Birth not used

Note that there will be rare cases where women will have had a stillbirth in the past by caesarean delivery where the previous stillbirth will not be captured in the data because it only includes previous live births. This could cause misclassification of the number of previous vaginal births but would not affect the number of previous caesarean deliveries. This rare event is not expected to impact on estimates of rates of uterine rupture in this study.

### Primary study factors

1. The number of previous caesarean deliveries
2. Whether or not there were any previous vaginal birth(s)

### Primary outcome

The primary outcome is uterine rupture, which is defined in the US Centres for Disease Control and Prevention (CDC) dataset as a full thickness rupture of the uterine wall and serosa. It does not include uterine dehiscence in which the fetus, placenta and umbilical cord remain contained within the uterine cavity; nor does it include silent, incomplete or asymptomatic separation.

The rates of uterine rupture will be reported as numbers and percentages and compared using adjusted odds ratios (95% confidence intervals).

### Secondary outcomes

All secondary outcomes are categorical and will be reported as numbers and percentages. Secondary outcomes are as follows -

#### Maternal

- Unplanned hysterectomy
- Blood transfusion
- Intensive care unit admission

#### Fetal

- Intrapartum fetal death and neonatal death (reported separately and as a combined total)
- 5-minute Apgar score < 4
- Neonatal seizures or serious neurological dysfunction (severe alteration of alertness excluding lethargy or hypotonia in the absence of other neurological findings)
- Admission to neonatal intensive care unit
- Neonatal ventilation immediately after birth
- Neonatal ventilation for more than 6 hours

While uterine rupture may lie on the causal pathway between the exposure and these secondary outcomes, no formal mediation analysis is planned.

### Variables

Continuous variables will be described using means and standard deviations (SDs) if normally distributed; otherwise median and interquartile range (IQR) will be used. Categorical variables will be reported as numbers and percentages (‘n (%)’ in the bullet list below). Demographic and clinical characteristics will be reported as follows:

- Total number of previous caesarean deliveries (3 categories; one, two, or three)
- Previous vaginal birth(s) (dichotomous; yes/no) - n(%)
- Labour type (dichotomous; spontaneous, induced/augmented)
- Maternal body mass index (BMI) at delivery (mean/SD)
- Maternal height in cm (mean/SD)
- Maternal age in years (median/IQR)
- Gestational age at birth in weeks (median/IQR)
- Birthweight in grams (mean/SD)
- Maternal race (4 categories; White, Black, American Indigenous or Alaskan Native, and ‘Other’) - n(%)
- Maternal cigarette smoking (dichotomous; yes/no) - n(%)
- Year of delivery (birth) of baby (categorical by year) - n(%)
- Payment type (dichotomous; public/private)
- Education (5 categories: (i) 12th grade or less; (ii) High school graduate, Tertiary credit or Associate degree; (iii) Bachelor’s degree; (iv) Master’s degree; (v) PhD or Professional degree) - n(%)
- Diabetes in pregnancy (3 categories: no diabetes, gestational diabetes, pre-existing diabetes) - n(%)
- Hypertension in pregnancy (dichotomous; yes/no) - n(%)
- Trimester of onset of maternity care (4 categories: first, second or third trimester; or no antenatal care)
- All secondary outcomes (listed in the previous section) are dichotomous (yes/no) variables

### Sample size

no sample size calculation will be performed. The maximum amount of available data will be used to maintain narrow confidence intervals, as uterine rupture is an uncommon event.

### Data collection and methods

Publicly available data from the CDC will be used and can be downloaded from https://www.cdc.gov/nchs/data_access/vitalstatsonline.htm Variables that are a planned part of the analysis will be interrogated to describe any missingness and extreme outliers.

### Analysis plan

Data will be downloaded from the CDC website https://www.cdc.gov/nchs/data_access/vitalstatsonline.htm for birth years 2011 to the latest available year.

If data missingness is present for the explanatory variables for more than 5% and less than 40% of eligible births then multiple imputation will be considered. Potential mechanisms of missingness and the relationship of missingness to the outcome and covariates will be assessed when deciding to conduct multiple imputation or a complete case analysis.

If multiple imputation is required, we will use fully conditional specification, with five imputed datasets. Any multiple imputation will include all explanatory and outcome variables, any spline terms, and derived terms including BMI, squared terms used to assess for linearity, and interaction terms (number of previous caesarean deliveries x previous vaginal birth(s)). Interdependencies amongst variables will be handled with the passive imputation method.

In the most recent available year of data, a preliminary analysis of continuous explanatory variables with be undertaken to assess their form in the multivariable analysis. The mean value of each 5%ile group will be plotted against the log-odds of uterine rupture in that group with Wilson confidence intervals. If there is an approximately linear relationship then the raw value will be used, if there is an approximate quadratic relationship, a centred squared term will be added to the base model in the backwards regression, and otherwise clinically relevant categorisation or spline terms will be considered.

Backward stepwise elimination will be conducted using log-likelihood ratio tests on the contribution of each explanatory variable remaining in the model. Variables will be eliminated one at a time if (1) they have the highest p-value; and (2) the p-value is > 0.05. If multiple imputation has been used, p-values for the likelihood ratio test in each imputed dataset will be pooled using the median p-value rule.

The following variables will be included in the base model as potential predictors of uterine rupture:

- Number of previous caesarean deliveries (not eligible for exclusion in the stepwise elimination)
- Previous vaginal birth(s) (not eligible for exclusion in the stepwise elimination)
- Labour induced or augmented versus spontaneous onset of labour (not eligible for exclusion in the stepwise elimination)
- Maternal age and (if supported in preliminary analysis) maternal age squared
- Gestational age (if the relationship is not linear, alternative forms will be explored)
- Maternal height and (if supported in preliminary analysis) maternal height squared
- Birthweight and (if supported in preliminary analysis) birthweight squared
- Maternal race
- Cigarette smoking
- Year of birth
- Payment type
- Trimester of onset of maternity care
- Interaction terms of the number of previous caesarean deliveries x previous vaginal birth(s)
- BMI and (if supported in preliminary analysis) BMI squared
- Hypertension in pregnancy
- Diabetes
- Education

Using the final multivariate fitted model, predicted probabilities of uterine rupture and the upper limits of the 95% confidence intervals will be tabulated by number of previous caesarean deliveries, previous vaginal birth, and labour induced or augmented versus spontaneous onset of labour.

#### Sensitivity analyses

If multiple imputation is used, the final multivariate fitted model from the imputed datasets will be replicated in the complete data for comparison.

#### Secondary outcomes

Among women with a ruptured uterus, secondary outcomes will be compared by the number of previous caesarean deliveries and previous vaginal birth(s) using Χ^2^ tests. If there is no association between number of previous caesarean deliveries or presence of a previous vaginal birth and the secondary outcome, the data will be reported by uterine rupture and no uterine rupture. If the outcome is associated with the number of previous caesarean deliveries or previous vaginal birth(s) then rates of the secondary outcome will be reported by number of previous caesarean deliveries or presence of previous vaginal birth(s) and whether uterine rupture occurred.

## Summary

This study will analyse more than 10 years of perinatal data from the US to assess the safety of TOLAC after one, two or three previous caesarean deliveries. Groups will be standardised for a minimum 21-month interpregnancy interval because prior to 21 months, shorter interpregnancy interval is associated with uterine rupture. Estimates for the predicted probability of uterine rupture by number of previous caesarean deliveries and whether there was a previous vaginal birth will be provided from the fitted models to support clinical decision-making.

## Data Availability

Publicly available data from the CDC will be used and can be downloaded from https://www.cdc.gov/nchs/data_access/vitalstatsonline.htm

## Notes

### Competing Interest Statement

The authors have declared no competing interest.

